# Experiences of Personalized Dementia Risk Education: A Qualitative Study to Refine the TEACH (Tailored Education for Aging and Cognitive Health) Behavioral Intervention

**DOI:** 10.64898/2026.01.27.26344961

**Authors:** L.E. Korthauer, A. De la Roca, R.K. Rosen, I. Arias, G. Tremont, J.D. Davis

## Abstract

**Background:** This study used qualitative methods to test and refine a framework for educating cognitively unimpaired individuals about their individual risk for Alzheimer’s disease and related dementias (ADRD) and intrapersonal health belief factors as part of the TEACH (Tailored Education for Aging and Cognitive Health) intervention.

**Method:** We assessed individuals’ ADRD risk factors and health belief concepts. Personalized data were presented individually, followed by a semi-structured phenomenographic interview. Applied thematic analysis was used to identify representative statements, trends, and differences.

**Results:** In N=11 individual interviews with middle-aged and older participants (ages 49-69; 45% women), participants had generally positive experiences of learning their personal dementia risk; the information was perceived to be unsurprising and occasionally consoling. They demonstrated a good understanding of the health belief concepts, including identifying relationships between intrapersonal health beliefs and health behaviors. Participants provided feedback on the visual aids and methods of conveying health belief information.

**Conclusions:** We used qualitative data from individual interviews to refine an explanatory framework for educating individuals about their personalized risk for ADRD and intrapersonal health beliefs that may be barriers or facilitators of health behavior change. The refined TEACH intervention is designed to promote long-term maintenance of target health behaviors in middle-aged adults to mitigate ADRD risk.

## Introduction

Fourteen modifiable risk factors account for approximately 45% of dementia cases globally^1^, making modification of health behaviors a public health priority. Middle age is a critical window for intervention, as Alzheimer’s disease and related dementia (ADRD) neuropathologies begin to accumulate 10-15 years or more before the onset of symptoms^2,3^. Generating sustained health behavior change during the midlife to early late life period could prevent or delay the onset of ADRD, resulting in greater quality of life and lower healthcare costs for the aging U.S. population.

Large, epidemiological or cohort studies have demonstrated that positive health behaviors such as regular physical activity, appropriate nutrition, cognitive activity, and social activity can mitigate dementia risk^4^. However, there have been relatively few randomized controlled trials aimed at primary prevention of dementia. One of the best-known studies, the Finnish Geriatric Intervention Study to Prevent Cognitive Impairment and Disability (FINGER), was a multi-component intervention designed for older adults at elevated risk of ADRD that included physical activity, cognitive activity, dietary modification, and vascular disease management^5^. Study outcomes showed that after two years, the multi-domain intervention resulted in 25% greater improvement on a cognitive composite measure compared to the control condition^6^. These findings were recently replicated in the U.S. POINTER study, which compared structured versus self-guided lifestyle intervention^7^. After two years, participants randomized to the structured intervention had significantly larger increases in a global cognitive composite than those in the self-guided group^8^.

As predicted, the U.S. POINTER and FINGER interventions increased the frequency of target health behaviors (e.g., adherence to dietary recommendations and physical activity guidelines). However, adherence to these interventions has been mixed. In the FINGER trial, overall adherence to the trial protocol was low, with only 19% of participants adhering to all components of the intervention (defined as 66% session attendance)^9^. Those who were non-adherent or minimally adherent had poorer cognitive outcomes^10^. The U.S. POINTER study had higher adherence in terms of session attendance (91% attendance for the structured intervention and 94.8% for the self-guided intervention), but adherence to the target health behaviors has not been reported. These findings highlight the importance of structured interventions to promote health behavior change but raise broader questions about the durability of treatment effects in multi-domain behavioral interventions after the cessation of active treatment.

Developing interventions based on principles from the science of behavior change is a way to increase long-term adherence to key health behaviors. Toward this end, we have begun developing a novel, multi-domain behavioral intervention targeted at health promotion in middle age: TEACH (Tailored Education for Aging and Cognitive Health)^11^. The TEACH intervention is theoretically grounded in the Health Belief Model, which identifies perceived threat of disease, perceived benefits and barriers to change, and self-efficacy as mediators of health behavior change^12^. TEACH is a personalized intervention designed to educate participants about 1) their personal risk for ADRD, including modifiable and non-modifiable risk factors and 2) intrapersonal health belief factors that are possible barriers or facilitators of health behavior change. These intrapersonal health belief factors were identified through the Science of Behavior Change Research Network^13^ and include seven constructs important for engagement of health behavior: future time perspective, executive control, response inhibition, self-efficacy, consideration of future consequences, deferment of gratification, and delay discounting (Table 1).

**Table 1.** Description of health belief factors.

| **Construct** | **Layperson-Friendly Title** | **Description** |
| --- | --- | --- |
| Future time perspective | Imagining your future | How much time you believe you have left in life and how you want to spend that time. |
| Response inhibition | Using your filter | Your brain's ability to manage impulses. |
| Delay discounting | Controlled reward | How much you want something right now, even if you could hold off for more later on. |
| Executive control | Staying on task | How well your brain can focus on a goal while ignoring distractions |
| Deferment of gratification | Delaying gratification | Your ability to put off something you want now to wait for something even better in the future. |
| Consideration of future consequences | Now and later | How you think about the long-term consequences of your actions. |
| Self-efficacy | Self-confidence | Your belief in your ability to reach a goal. |

*Note.* Descriptions were written for ~6<sup>th</sup> grade reading level, per NIH recommendations (Fleisch-Kincaid grade level = 6.2).

Although the Health Belief Model has only rarely been applied to dementia prevention, studies from our group and others have demonstrated that these health belief constructs are associated with lower ADRD risk and greater engagement in brain health-relevant behaviors^14,15^. However, to our knowledge, no prior studies have used these intrapersonal health belief processes as an active component of a behavioral intervention. This is a major gap, as increasing individuals’ insight into these intrapersonal processes can help them understand not just *what* to do to lower ADRD risk, but *why* and *how* to make sustained behavioral changes. TEACH is expected to enhance an existing health education program by providing personalized education about individual ADRD risk factors and intrapersonal health belief processes. Our intervention addresses five components that have been identified as critical for maintenance, not merely initiation, of behavior change: motivation, self-regulation, resources (psychological and physical), habits, and environmental and social influences^16^.

In our initial development of the TEACH intervention, we conducted focus groups to develop an explanatory framework for educating participants about ADRD risk and intrapersonal health belief factors^17^. Participants were presented with health information about a fictitious person, including her risk for ADRD and her personal levels of each health belief trait. Participants generally understood the health belief constructs and described subtle distinctions between related concepts. They were able to describe links between health belief factors and health behaviors (e.g., diet, physical activity). Most participants were enthusiastic about the prospect of learning their own personal health information. We used this information to develop a framework for educating individuals about their ADRD risk and health belief profile, including layperson-friendly descriptions, graphics, and a protocol for a clinician to guide the education session.

The next step in this NIH Stage I behavioral intervention development study was to use qualitative interviews to test this explanatory framework by educating individuals about their personal ADRD risk and health belief profile. This component of the study used a phenomenographic approach^18,19^ which is designed to collect qualitative data about participants’ experiences of learning this information. Our interviews focused on participants conceptions and understanding of the elements in their ADRD risk profile and health beliefs. We used this qualitative data to assess the understandability, appropriateness, and applicability of this information to individuals’ engagement in relevant health behaviors. Data from this step will be used to further refine the TEACH intervention in preparation for a pilot randomized controlled trial to examine the feasibility of delivering the TEACH intervention and provide preliminary estimates of its efficacy compared to basic health education alone.

## Methods

### Participants

Participants were recruited through the general Rhode Island community, including outreach through the Rhode Island Alzheimer’s Disease Prevention Registry and the Rhode Island Hospital Neuropsychology Program. Participants met the following inclusion criteria: a) age 45-69 years; b) cognitively unimpaired status (Minnesota Cognitive Acuity Scale^20^ > 52), and c) written and oral English language fluency. Of 109 participants pre-screened for eligibility, 58 were contacted and 17 responded to outreach attempts. Twelve of those 17 individuals completed telephone screening and scheduled a study visit; 1 participant cancelled their visit and declined to reschedule, arriving at a final analytic sample of N=11 participants. All study procedures were approved by the Brown University Health institutional review board.

### TEACH ADRD Risk and Health Belief Assessment

Complete details of the TEACH study protocol are reported elsewhere^11^. Participants completed the TEACH assessment, which consists of seven instruments from the Science of Behavior Change Research Network, the Australian National University-Alzheimer’s Disease Risk Index (ANU-ADRI)^21^, and the Pittsburgh Sleep Quality Index^22^. The SOBC Research Network measures included the Future Time Perspective Scale^23^, the Deferment of Gratification Scale^24^, the Consideration of Future Consequences Scale^25^, the Generalized Self-Efficacy Scale^26^, the Monetary Choice Task (a measure of reward sensitivity/delay discounting)^27^, Parametric Go-No Go Task (response inhibition)^28^, and the Attentional Network Test (flanker interference subtest; executive control)^29^. Self-response measures were administered via computerized RedCap survey, while neurocognitive measures (Parametric Go-No Go and Attentional Network Test) were administered on a laptop computer using e-Prime software (Psychology Software Tools).

A graphic was created for each participant illustrating their personal risk for ADRD (Figure 1A), based on their responses to the ANU-ADRI and other self-report measures. This graphic was developed during the initial focus group phase of the TEACH development project^17^. The graphic includes both non-modifiable risk factors (age, gender, family history, early life education) and modifiable risk factors (alcohol use, body weight, cholesterol, cognitive activity, diabetes status, diet, mood, pesticide exposure, physical activity, smoking status, social network, traumatic brain injury).

**Figure 1.**
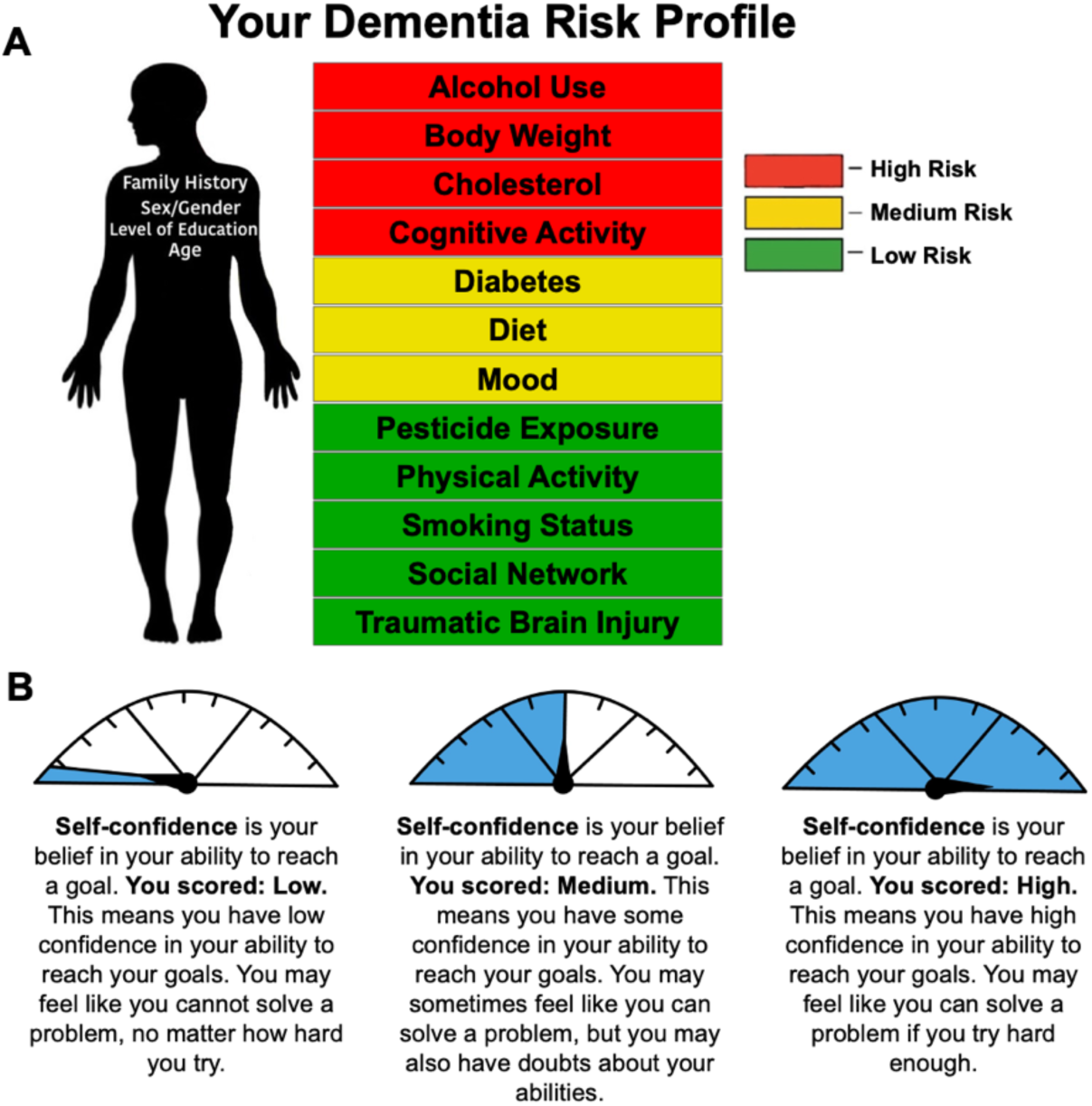
A) Graphic showing modifiable (red, yellow, green) and non-modifiable (factors listed on the person’s body) risk factors for Alzheimer’s disease and related dementias (ADRD). Participants’ images were personalized based on their responses to the TEACH assessment measures. B) Example of the speedometer icon and descriptions for a health belief trait. The health belief profile included graphics and descriptions for each of the seven traits.

A health trait profile was also created for each individual based on their responses to the seven SOBC Research Network measures. These scores were determined based on comparison to a normative dataset collected in our prior developmental work^14^. Each score was labeled “low” (>1 SD below the mean), “average” (±1 SD from the mean), or “high” (>1 SD above the mean). An image of a speedometer was accompanied by a brief, layperson-friendly description of the construct and interpretation of the score (Figure 1B); these were also developed during the initial focus groups.

Based on feedback from those focus groups, in the interviews we presented the health belief traits in two ways. One health belief profile was organized by domain (“How You View the Future” [future time perspective], “Your Ability to Control Your Behavior” [response inhibition, executive control], “Your Confidence In Your Ability to Change” [self-efficacy], and “How You Consider the Future When Making Decisions” [delayed gratification, consideration of future consequences, delay discounting]). A second health belief profile, containing the same descriptions and scores, was organized by strengths (“Traits that May Make It Easier to Make a Health Change” [average or high scores] and weaknesses (“Traits That May Make It Harder to Make a Health Change” [low scores]).

### Phenomenographic Qualitative Interviews

The ADRD risk profile and health belief profile were presented to each individual by a clinical neuropsychologist (JD or LK) following a standardized protocol. Participants were first shown their ADRD risk profile and educated about their non-modifiable and modifiable risk factors. Information was provided about how the scores were derived. The interpretation of each risk factor (green = low risk, yellow = medium risk, red = high risk) was discussed. Qualitative interviews were then performed following a standard discussion guide developed using a phenomenographic approach.^18,30,31^ The discussion guide included questions to gauge reactions to the presentation of information, assess understanding of the information (e.g., what risk factors a person should work on most, what it means for a risk factor to appear in green versus yellow), perceptions of ability to change the risk factors, and perceived relevance of the information for a person’s health. Probes were used to explore responses and seek clarification.

Participants were then shown their personal health belief profile. Order of presentation (clustered based on content area versus strengths and weaknesses) was counterbalanced across participants to minimize novelty or familiarity effects on reactions to the information. Participants were educated about their personal levels of each health belief factor. Discussion questions gauged their understanding of the content, perceived applicability of the health beliefs to their own health behaviors, and their overall reactions to the explanatory process. Participants were asked to identify any constructs that were confusing, redundant, irrelevant, or seemed like an inaccurate representation of themselves. They were then shown the second health belief profile, clustered in a different way, to assess whether method of presentation influenced understanding and motivation to change.

### Qualitative Analysis

Each interview was digitally recorded and professionally transcribed. Identifiers were removed from the transcripts. Transcripts and field notes were reviewed by two members of the research team with qualitative methods expertise (RKR, GT). A codebook of both deductive and inductive codes was created. Deductive codes focused on components of each image, responses to the presented constructs, and the comparison of the two versions of the health belief profile. Inductive codes included feedback about the experience of learning their ADRD risk, advice sought from facilitators (tracked as potential indicators of sources of confusion or needed additional information) and recommended changes to the images. The coders individually reviewed the transcripts, then met to discuss and agree-upon coding. Agreed-upon codes were entered into NVivo software (version 14, 2023). Code summaries were written and then were reviewed and discussed with the entire study team.

## Results

We conducted 11 qualitative interviews (*M* age = 58.8 years, range = [49-69]; 45% women and 45% men (1 preferred not to answer); 91% non-Hispanic White; *M* education = 15.1 years, range [12-18]).

Overall, the interviews demonstrated that the ADRD risk information and health belief constructs were understandable and relevant to non-expert individuals. Most participants found the experience of learning their personal ADRD risk level to be acceptable and unsurprising, with some finding the experience consoling. They found the graphics used to display ADRD risk to be clear and understandable. Several spontaneously expressed a desire for ADRD risk information to be accompanied by specific recommendations for risk reduction. Many expressed that the information about health beliefs felt like it confirmed something they already knew about themselves. Some participants wished for a greater amount of detail about their specific scores, but this was a minority opinion.

### ADRD Risk Factor Presentation

Individual participants’ ADRD risk factors were presented graphically (Figure 1A) with accompanying information provided by a clinical neuropsychologist. Most participants expressed understanding of the risk factors and found the graphics helpful. One participant expressed confusion about the classification of modifiable versus non-modifiable risk factors, specifically educational attainment as a non-modifiable risk factor.

> *“I think the level of education kinda confuses me, because it is something that you do have an effect on, sort of…I guess that kinda threw me off, because it was like, yeah, I can’t change my family history. I can’t change my gender, but it’s like no, I made a decision to educate myself”* [Participant 02].

After receiving education about their personalized ADRD risk, many participants said that they already knew the information presented: *“Honestly, not surprising”* [03] and “*Nothing’s a super surprise”* [04]. While many participants stated that their risk factor information was unsurprising, several also said that they learned something new about ADRD risk. Risk conferred by pesticide exposure was specifically noted to be new information for some participants: “*Pesticide exposure wasn’t even on my radar to think about. Like I know they’re terrible and all sorts of things, and cancers and stuff, but wasn’t really on my radar for cognitive decline”* [02]. Others reflected that they knew about some risk factors for overall health, such as diabetes or high cholesterol, but they did not realize that those factors specifically confer risk for ADRD. Several participants commented that receiving the risk information itself was valuable, but they wanted more information about what to do to mitigate risk:

> Participant 10: *“It makes me feel like, ‘How am I going to get – how am I going to achieve this?”*
>
> Interviewer: *“So…when you see everything presented this way, it feels maybe a little overwhelming…kind of like, ‘Whoa, there’s a lot I have to do’?”*
>
> Participant 10: *“No, because you know what? It’s not surprising, so it doesn’t overwhelm me…but it’s like, ‘I don’t know how to correct it.’”* [participant continues to discuss wanting more information to obtain skills needed to address ADRD risk factors].

Another participant commented: “*I want you to tell me exactly what three things to do to avoid Alzheimer’s, and I’ll do exactly what you say”* [01].

Regarding the ADRD risk education process itself, participants overall had positive reactions to the way in which information was disclosed: *“It felt collaborative…we were sharing information back and forth…it felt supportive”* [04]. No participants, even those with multiple ADRD risk factors, expressed an adverse reaction to the ADRD risk education. During the discussion, participants often narratively explained why they thought they received the scores they did, providing examples from their lives, experiences, and known medical issues (e.g., high cholesterol, alcohol use, ADHD, etc.).

The interviewers also solicited feedback about the image used to display a person’s level of each ADRD risk factor. Reactions were positive overall, with participants expressing, *“The graph…says it all,*” *“This felt very clear,”* and *“It all just makes sense to me*” [10]. One participant said:

> *“I like the way it’s presented, because I think it kind of jumps at you…what are the things that you need to consider changing, and what are the things you’re already doing right, so you’re getting some positive reinforcement along with the…okay, but here’s some stuff that we probably could work on”* [02].

One participant wanted more detail about how much risk each factor color conferred: “*it would be interesting if it was on the scale of 1 to 10…because it’s sort of stark. You go from yellow to red, you might be on the borderline”* and felt that the non-modifiable risk factors on the person’s body *“almost don’t get enough play”* [09]. However, this was not a majority opinion. Most other participants noted that the color coding made the content clear and understandable: “*It’s an effective visual aid*” [06].

### Health Belief Education

As with the ADRD risk education, most participants found the discussion of their personal health belief traits to be a positive experience and largely unsurprising. Most participants agreed when asked, or said directly, that the information presented felt like it was confirming something they already knew about themselves or that it felt “*pretty true*” [02] to them. For example, one participant stated: “*I would have to say that I’m not necessarily sure that I’ve learned anything new. I think it just kind of…reaffirms what I already know”* [10]. Another participant expressed that the process of being educated about personal health beliefs felt true to their inner representation of themself: [Participant 05]: *“It’s…like a layer’s been taken off. It’s like – so you can see – you can see who I am.*” [Interviewer]: “*And do you feel like I got it right?”* [Participant 05]: “*Yeah.*”

Although most participants found the information to largely fit their prior conceptions of themselves, some expressed surprise about a piece of information that differed from their sense of self. For example, one participant reflected, “*I would have thought that my self-confidence average was a little bit…higher…I’m not surprised at all about staying on task, in fact I’m a little bit surprised it’s average* [participant expected it to be lower]” [04]. Another had a reaction to their low score on delay discounting:

> “*It’s the only one I really disagree with…I usually wait on everything, you know?…Just this morning, I was like, ‘I got to get a new pair of boots for the winter.’ And I said, ‘Nope. Not going to spend the money.’…I don’t feel that one matches”* [10].

When a person did express disagreement with an element of their health belief profile, it was typically restricted to one or two scores.

### Health Beliefs and Engagement in Health Behaviors

Many participants drew connections between their health belief traits and everyday behaviors. They provided examples regarding a range of health behaviors, including physical activity, dietary choices, alcohol use, and cognitive activity. Each participant had a different set of traits that they considered the greatest barriers and facilitators of behavior change. For example, one participant reflected about the following health beliefs related to their alcohol consumption:

> “*Consideration of future consequences…I don’t want to have a headache, I don’t want to be sick, so I’m not going to have any more…[and, regarding other important traits] I think self-confidence and just using my filter”* [05].

Several people also reflected that how a particular health belief affects behavior may depend on context.

> *[Regarding response inhibition] “I was sort of inhibited socially, so on those moments where I decided to be spontaneous and go with the impulse, you know, it could be a reward…[In contrast], alcohol and smoking. Well, that’s the more typical associations with impulse control, which I’ve had troubles with those, too”* [09].

One health belief trait that elicited differing reactions among participants was future time perspective. Individual responses to this concept varied, with some people feeling like having a high future time perspective score would motivate them to change health behaviors (because of having more time to make a positive change) and others expressing that a high score would be demotivating (because engaging in healthier behaviors would feel like a less enjoyable use of that remaining time).

> “*You know, it’s like…if I’ve got 20 years left, what do I want to spend the 20 years on? It sure as hell isn’t making sure I fit into a size four again”* [01].
>
> *“I need to be concerned about keeping my mind as – as strong as possible. Or as healthy as possible. So…considering my future… I need to be able to stay sharp, as sharp as possible”* [10].

Overall, no specific traits or combinations of traits were identified as the most important barriers or facilitators of health behavior change. When asked whether any concepts relevant to health behavior were missing from their profile, several participants cited the concept of grit or determination (e.g., “*disciplined*” [05], “*sheer determination*” [08], “*bullheadedness*” [06]) as absent.

## Discussion

This study used a phenomenographic approach to examine participants’ ways of understanding and experiencing a structured approach to learning about personalized ADRD risk and health belief factors that may serve as barriers or facilitators of health behavior change^17^. Our findings demonstrate the feasibility, acceptability, and appropriateness of this approach for non-expert, middle-aged adults. Importantly, the personalized approach to communicating ADRD risk, including both modifiable and non-modifiable factors, was well received and did not elicit distress, even among people with numerous risk factors. This study represents a critical step in the iterative design process for our TEACH intervention, which aims to promote long-term maintenance of health behaviors for primary prevention of ADRD.

The goal of phenomenography is to examine variations in conceptions of, and the process of learning, new information^18,30^, in this case personal information about ADRD risk and factors important for risk reduction. Thus, our approach emphasized the process and experience of learning personalized information, allowing us to understand how to communicate in a way to facilitate understandability, minimize distress, and to identify what challenges may arise. We observed different levels of engagement with the presented information, from passive receipt to active integration, with many participants drawing connections between health beliefs and specific health behaviors.

Prior studies examining health education in the context of risk for ADRD have primarily focused on disclosure of biological risk factors, including *APOE* status or AD biomarker information. For example, the REVEAL study^32^ found that disclosing participants’ *APOE* status did not result in short-term psychological risks, and a majority of participants (63%) recalled the information one year later.^33^ Findings are similar for disclosure of AD biomarkers such as amyloid positivity, with the IMPACT-AD study finding that 82% experienced positive feelings about the disclosure process^34^. Importantly, disclosure of personalized ADRD risk or disease biomarker information seems to be a motivating factor for health behavior change, including increased exercise and cognitive activity^34,35^, though other studies have reported minimal behavior changes post-disclosure.^36,37^ In this study, we educated participants about multiple modifiable and non-modifiable risk factors, including medical factors (e.g., hypertension, high cholesterol), sensory changes (e.g., hearing impairment), specific behaviors (e.g., physical activity level, social activity), and environmental exposures. Participants largely understood the information presented, though some acknowledged that they had not previously considered these factors in the context of ADRD risk specifically. While many participants felt that the ADRD risk education confirmed information they already knew, this did not diminish the perceived value of the experience. The desire for next steps and concrete guidance on how to proceed was a consistent theme. This highlights the importance of the TEACH intervention itself, which will provide evidence-based recommendations for each of the covered risk factors.

Our findings also speak to the acceptability and appropriateness of the health belief constructs. Though each of these constructs has previously been demonstrated to mediate or moderate health behavior change, they have rarely been formally incorporated into an ADRD prevention intervention. The health belief traits are psychological factors that are abstract, making it critical to ensure participants’ understanding of their relevance for everyday behavioral decisions. Thus, our earlier intervention development work focused on eliciting words and phrases from participants to describe these concepts. In the current study, most participants felt that their individualized profiles were understandable and aligned with their self-concept. Where discrepancies arose, they were typically limited to one or two traits and did not appear to undermine overall acceptability of the health belief framework. The visual aids, particularly color-coded graphics, were viewed as highly effective. This promotes the feasibility and fidelity of information delivery, particularly in lower health literacy populations^38^.

The discussion of health belief traits also revealed complex and sometimes contradictory interpretations of specific health beliefs, particularly regarding future time perspective. Consistent with our formative work with focus groups^17^ and prior literature^39^, some participants viewed a longer future time perspective as motivational, while others felt that it reduced their sense of urgency to make health behavior changes. This variability speaks to the role of an individual’s prior experiences and personal context in shaping understanding of the health belief constructs^39,40^. This does not necessarily undermine the potential efficacy of future time perspective as a motivator of health behavior change. However, it highlights a need for interventionists to flexibly tailor the intervention to help participants build awareness of these health traits, whether a given trait is perceived as a barrier or facilitator of health behavior change.

Using the data from this phenomenographic analysis of individual interviews, we plan to make minor changes to our structured ADRD risk education protocol in preparation for a randomized controlled trial of the TEACH intervention. This includes making clearer distinctions between early life education as a non-modifiable risk factor versus cognitive activity level as a modifiable risk factor in mid- to late-life. We will also provide an opportunity for participants to reflect on which health belief factors feel more or less relevant to them for health behavior change. This adds another aspect of tailoring for the intervention by remaining sensitive to person-level contextual factors, rather than taking a “one-size-fits-all” approach to connecting health beliefs to health behavior change. Our plan is to follow this protocol for ADRD risk and health belief education prior to beginning the TEACH intervention. The TEACH intervention provides evidence-based recommendations about modifiable ADRD risk factors, addressing participants’ desire for actionable steps to guide behavior change. TEACH also incorporates intrapersonal health belief factors through structured goal-setting exercises to identify behavior change targets, anticipate barriers, and increase awareness of intrapersonal factors that promote long-term behavioral maintenance. To incorporate the concepts of “grit” or “determination” that some participants felt were missing from the health belief profile, we have added content to the TEACH intervention, including 1) a values exploration exercise (“finding your why”) to connect short-term behavioral goals to underlying values; and 2) psychoeducation regarding lapse prevention and how to cultivate long-term resilience when making health behavior changes.

In summary, qualitative analysis of individual interviews demonstrated the feasibility and acceptability of educating middle-aged adults about their personal risk for ADRD and health belief factors that may promote health behavior change. Individuals found the health belief information to be understandable, relevant, and largely affirming of their self-perceptions Our findings underscore the importance of coupling education about ADRD risk with actionable guidance, which may promote long-term maintenance of target health behaviors.

## Acknowledgments

We would like to acknowledge support of the Biostatistics, Epidemiology, and Research Design service core from Advance Clinical and Translational Research (Advance RI-CTR), which is funded by Institution Development Award Number U54GM115677 from the National Institute of General Medical Sciences of the National Institutes of Health. The content is solely the responsibility of the authors and does not necessarily represent the official views of the National Institutes of Health.

## Data Availability Statement

The data are not publicly available due to the nature of the qualitative data and risk that even de-identified transcripts could contain information that could compromise the privacy of research participants.

## Ethical Approval and Informed Consent Statements

All study procedures were approved by the Brown University Health institutional review board. All participants provided written informed consent.

